# Intertransverse process block for chronic postsurgical pain in cardiac surgery: protocol for a double-blinded randomized controlled trial

**DOI:** 10.1101/2025.07.11.25331342

**Authors:** Henry Man Kin Wong, Ranjith Kumar Sivakumar, Wai Tat Wong, Albert Kam Ming Chan, Zion Ho Sum Yeung, Pik Yu Chen, Sherry Tsz Wai Tang, Mandy Hiu Man Chu, Randolph Hung Leung Wong, Kwok Ming Ho

## Abstract

**Introduction:** Chronic postsurgical pain (CPSP) after cardiac surgery is a significant concern. Despite the known association between acute pain and CPSP, advanced pain management strategies have not reduced its incidence. Preventing CPSP requires optimizing acute pain control and disrupting central sensitization. The side effects and risks associated with chronic use of current opioid-based cardiac anesthesia necessitate the adoption of multimodal analgesia. Regional anesthesia is a promising alternative, though existing techniques in cardiac surgery have notable limitations. The intertransverse process block (ITPB) is a novel regional technique that offers potential somatic and visceral analgesia. Recent studies demonstrate consistent local anesthetic spread to the intercostal, paravertebral, and epidural spaces, suggesting broader pain control. ITPB may provide a simpler, safer approach in cardiac surgery, reducing the risks of pleural puncture and bleeding. We hypothesize that ITPB will improve pain control, quality of recovery, and health-related quality of life, thereby mitigating chronic postsurgical pain.

**Methods:** This is a single-center, randomized, double-blinded, placebo-controlled trial with intention-to-treat analysis. Elective patients awaiting coronary artery bypass grafting, with or without valvular repair or replacement, will be recruited. Ninety-six participants will be randomly assigned to either ITPB or control group. The ITPB group will receive bilateral ITPBs with 20 ml 0.25% levobupivacaine on each side at the T4-5 level under ultrasound guidance, administered before anesthesia induction. Sham blocks, with equal volume of normal saline, will be performed in the control group. The primary outcome is the quality of recovery, assessed using the 15-item Quality of Recovery questionnaire, at 24 hours after tracheal extubation. Secondary outcomes include Numerical Rating Scale pain scores, patient satisfaction, and opioid consumption within 72 hours post-extubation, duration of mechanical ventilation, length of stay in the ICU and hospital, and opioid-related side effects. The incidence of CPSP at 3, 6, and 12 months will be measured, along with pain interference via the Brief Pain Inventory and the Short-Form McGill Questionnaire-2.

**Discussion:** Current pain management strategies often rely heavily on opioids, which can have significant side effects and may not adequately address chronic postsurgical pain. This study investigates the efficacy of the intertransverse process block, a novel regional anesthesia technique, in reducing both acute and chronic postsurgical pain in cardiac surgery. Randomized controlled trials on intertransverse process block in cardiac surgery are limited. The results of this study will help define the role of intertransverse process block in reducing chronic postsurgical pain in cardiac surgical population.

**Clinical trial registration:** This trial has been prospectively registered at http://clinicaltrials.gov: NCT06946290

## Introduction

The cornerstone of modern pain management in cardiac surgery lies in proactively preventing the development of chronic postsurgical pain (CPSP). This paradigm shift emphasizes pre-emptive interventions to disrupt the cascade of events and central sensitization that transform acute postoperative pain into a persistent, debilitating condition [1–4]. CPSP, a significant and often underestimated consequence of cardiac surgery, affects a substantial proportion of patients, with prevalence rates ranging from 28% to 56% in the years following surgery [5–8]. Its impact extends beyond discomfort, significantly impairing daily function, reducing quality of life [8–9], and imposing a substantial economic burden on the healthcare system.

The pathophysiology of CPSP involves a complex interplay of factors, with central sensitization playing a pivotal role [3]. This process, characterized by heightened neuronal excitability within the central nervous system, amplifies pain signals and can lead to persistent pain even after the initial injury has healed. Therefore, a crucial aspect of CPSP prevention lies in disrupting the afferent nociceptive signals transmitted from injured tissues to the spinal cord and brain, thereby preventing the establishment and perpetuation of central sensitization [1,4].

While opioids have traditionally been the mainstay of postoperative pain management, their use is not without significant drawbacks. Opioids can have dose-dependent side effects, including respiratory depression, nausea, and constipation. Furthermore, prolonged opioid use can lead to tolerance, opioid-induced hyperalgesia, and an increased risk of chronic opioid dependence [10–12]. Recognizing these limitations, current pain management strategies emphasize multimodal analgesia, incorporating non-opioid medications and regional anesthesia techniques to optimize pain control while minimizing opioid-related risks.

Non-steroidal anti-inflammatory drugs (NSAIDs) offer a valuable adjunct in pain management, but their use in cardiac surgery is often limited due to concerns about bleeding complications and potential renal impairment. Other non-opioid analgesics, such as paracetamol and gabapentinoids, have shown limited efficacy in managing the intense pain associated with sternotomy [13]. Regional anesthesia techniques have emerged as promising strategies for both acute pain management and potential CPSP prevention in various surgical settings [14–17]. Techniques such as epidural anesthesia and paravertebral blocks have demonstrated efficacy in reducing postoperative pain intensity and opioid requirements [14–15]. However, their application in cardiac surgery presents unique challenges. Epidural anesthesia carries the risk of neuraxial haematoma due to systemic anticoagulation and heparinization, while paravertebral blocks may be associated with complications such as pneumothorax and pleural puncture [18]. Erector spinae plane block (ESPB) has shown inconsistent results in reducing postoperative pain and morphine consumption in cardiac surgery [19–24]. Parasternal plane blocks offer advantages over neuraxial techniques [25–26] but may not adequately address visceral pain.

While regional anesthesia is crucial for managing acute postoperative pain, its impact on CPSP remains largely unknown. The potential for regional anesthesia to reduce CPSP has been identified as one of the top research priorities in anesthesia and perioperative care [27]. Intertransverse process block (ITPB) is a novel and promising alternative, targeting the paravertebral space through extra-paravertebral injection within the intertransverse tissue complex, posterior to the superior costotransverse ligament (SCTL) [28–29]. Recent MRI studies have demonstrated consistent spread of local anesthetic to the ipsilateral intercostal, paravertebral spaces, neural foramina, and epidural space following ITPB [30], suggesting potential for both somatic and visceral analgesia. ITPB has shown preferential spread of local anesthetic to the epidural space and neural foramina compared to ESPB, and effective analgesia in breast and video-assisted thoracoscopic surgeries [31–32]. Compared to other regional techniques in cardiac surgery, ITPB may offer a simpler and safer approach with reduced risk of pleural puncture and bleeding. Therefore, this trial will assess the analgesic efficacy of ITPB in mitigating both acute and chronic postsurgical pain in cardiac surgical patients. We hypothesize that ITPB will reduce CPSP at 3 months after cardiac surgery, improve pain control, quality of recovery, and health-related quality of life.

## Materials and methods

### Study population and design

The protocol of this study follows the SPIRIT checklist. This is a single-centre, double-blinded, randomized controlled trial. Ethical approval was obtained from the Joint Chinese University of Hong Kong-New Territories East Cluster Research Ethics Committee on 22^nd^ May 2023 (CREC Ref No 2025.177-T). The study is registered at http://clinicaltrials.gov (NCT06946290), with the registration date of 23^rd^ April 2025. With an annual volume of approximately 400 elective cardiac cases, the recruitment is anticipated to start on 1^st^ November 2025 and to be completed by 31^st^ October 2026. Data collection will be completed by 31^st^ October 2027, and the results will be expected by 30^th^ December 2027. Eligible patients will be those undergoing elective cardiac surgery at Prince of Wales Hospital, a university teaching hospital with 1,650 beds in Hong Kong. All patients will be admitted to a 28-bed ICU for early postoperative care, monitored with 1:1 nursing at all times, and are expected to be discharged to a high-dependency cardiac ward within 24 hours after surgery.

### Randomization and concealment

Randomization will be performed in 1:1 ratio using the REDCap randomization module in the study database, allocating participants to either the ITPB (intervention) or control group in randomly permuted blocks of size four. Sequentially numbered, coded, sealed, opaque envelopes, each containing the group assignment of either interventional or control are then prepared by a third party who takes no further part in the study. The ITPB syringes will be prepared under strict aseptic conditions by a nurse not involved in the study, with blind labelling. The surgical team, blinded to group allocation, will perform standardized surgical procedures. Anesthesiologists and nurses responsible for data collection in both ICU and wards are also blinded to the treatment allocation.

### Eligibility criteria

Adult patients aged 18 years or older undergoing elective CABG, valve repair/replacement, or combined CABG/valve procedures via sternotomy will be included. Patients admitted for emergency surgery, aortic surgery, redo surgery, history of chronic pain or being on chronic opioids/sedatives, preoperative renal failure requiring renal replacement therapy or whose creatinine clearance <30 ml.min^-1^ (calculated by Cockcroft-Gault formula), liver dysfunction (liver enzymes twice upper limit normal), left ventricular ejection fraction <40%, requirement of mechanical hemodynamic support, intraoperative remifentanil use, and unable to provide informed consent will be excluded.

### Anesthesia and interventions

All patients will receive standard cardiac surgery monitoring. General anesthesia will be induced with midazolam (0.01-0.05 mg.kg^-1^), fentanyl (2-5 µg.kg^-1^), and rocuronium (0.5-1 mg.kg^-1^) to facilitate intubation. Anesthesia will be maintained with sevoflurane and propofol infusion, targeting a Bispectral Index of 40-60. ITPB will be performed after anesthesia induction, with the patient in the lateral decubitus position. Intraoperative opioids (fentanyl and morphine) will be administered at the discretion of the anesthesiologist. Postoperative analgesia will be identical in both groups, including patient-controlled analgesia (PCA) morphine (one milligram bolus, lockout period of five minutes, 4-hour maximum dose of 25 mg) for 72 hours after surgery, oral analgesics (paracetamol 1 g every six hours, dihydrocodeine 30 mg thrice a day), and on-demand antiemetics (intravenous ondansetron 4 mg every eight hours). Rescue analgesics may be prescribed as needed. Upon ICU admission, propofol infusion will be stopped to facilitate ventilator weaning using Adaptive Support Ventilation (ASV), which adjusts parameters based on the patent’s lung mechanics and effort. Pain will be assessed regularly in the ICU and on the ward. After extubation, pain scores will be assessed at 2, 4, 8, 12, 24, 48, and 72 hours. PCA morphine will be prescribed for patients with moderate to severe pain. Nausea, vomiting, and rescue antiemetics will be documented.

### Ultrasound block placement

The intervention group will receive bilateral ITPB prior to anesthesia induction, while the control group will receive sham blocks. All blocks will be performed by an experienced anesthesiologist who has completed more than fifty successful ITPB procedures, using a Philips EPIQ ultrasound system with a curved array transducer (C5-1) and 80 mm echogenic nerve block needle (SonoTAP; PAJUNK, Germany). ITPB will be performed with patients positioned in the lateral decubitus position. The target intervertebral level (T4-5) will be identified and marked using a preview ultrasound scan. The transducer will be placed two to three centimetres lateral to the spinous process. Under strict asepsis, a single-level (T4-5) ultrasound-guided ITPB will be performed using in-plane needle insertion from lateral to medial, targeting the medial aspect of the retro-SCTL space. Correct needle position will be confirmed by distension of the retro-SCTL space following a test bolus of 1-2 ml of 0.9% normal saline. Then, 20 ml of 0.25% levobupivacaine or 0.9% normal saline will be injected in small aliquots. The same procedure will be repeated on the contralateral side using the same volume of study medication. The time required to perform the block, from needle insertion to removal, will be recorded.

### Outcome measures

Although acute pain is recognized as an important predictor for the development of CPSP, different acute pain management strategies have failed to reduce the incidence of CPSP, suggesting a complex mechanistic link between acute and chronic pain after surgery. Evidence indicates that pain-related functional interference and patient-reported outcomes, such as quality of recovery, may be associated with the development of CPSP [33]. Therefore, the primary outcome of this study is the quality of recovery, measured by the 15-item Quality of Recovery questionnaire score (QoR-15) at 24 hours after tracheal extubation. QoR is recommended for assessing patient comfort after surgery and is a highly valid and reliable patient-centered outcome measure [34].

Secondary outcomes include Numerical Rating Scale (NRS) pain scores at 2, 4, 8, 12, 24, 48 and 72 hours after tracheal extubation, patient satisfaction with pain management, postoperative morphine consumption at the above time points, time to first morphine rescue (in minutes), intraoperative opioid consumption (converted into morphine equivalent doses), duration of mechanical ventilation, length of stay in ICU and hospital, opioid-related side effects, such as postoperative nausea and vomiting (PONV), incidence of CPSP at 3, 6, and 12 months, and pain interference assessed using the Short-Form McGill Pain Questionnaire-2 (SF-MPQ-2) and the Brief Pain Inventory (BPI) Interference Scale at 3, 6, and 12 months postoperatively.

CPSP is defined as persistent pain following surgery that was not present before the procedure or has different characteristics, with other possible causes of pain excluded. Participants reporting CPSP will be further assessed for severity and impact based on the recommendations from the Initiative on Methods, Measurement, and Pain Assessment in Clinical Trials (IMMPACT). This includes the SF-MPQ-2 to assess the sensory pain qualities and affective components, and the BPI to assess pain interference with physical functioning.

The Chinese version SF-MPQ-2 [35] will be used to measure sensory and affective aspects of pain. It evaluates chronic pain symptoms on an 11-point numerical rating scale (0 = none, 10 = worst possible). It includes three sensory descriptors and one affective descriptor. The four subscales will be calculated as a mean of items in each subscale, and the total score will be the mean of all 22 items. Higher scores indicate more intense symptoms. The Chinese version of the Brief Pain Inventory (BPI) Interference Scale [36] will be used to evaluate the extent to which pain interferes with various aspects of functioning, including physical and emotional functioning, and sleep.

### Data collection

After screening for eligibility, patients will receive an information sheet outlining the main aspects of the trial. The study will be discussed with a research nurse before written informed consent is obtained. All data will be collected by research team members who are blinded to group allocation. Patient demographics and body mass index will be recorded. Cumulative opioid consumption and time to first morphine rescue will be extracted from the PCA pump. At 2, 4, 8, 12, 24, 48, and 72 hours post-extubation, pain scores at rest and during coughing will be assessed using NRS from 0 to 10, where 0 indicates no pain and 10 points indicates the worst pain imaginable. Patients will also rate their overall satisfaction with pain management using a verbal analogue scale (0 = worst possible, 100 = best possible) at predefined time points. Nausea, vomiting, and the use of rescue antiemetics will be documented. The validated Chinese version of the QoR-15 questionnaire [37] will be administered at baseline (preoperatively) and at 24 and 72 hours postoperatively after tracheal extubation. The SF-MPQ-2 and BPI will be used to assess CPSP at 3, 6, and 12 months postoperatively.

The following medical and surgical data during the hospital stays will be extracted from electronic record:

1. Patient demographics (age, gender, EuroScore)
2. Type of surgery
3. Duration of surgery and duration of cardiopulmonary bypass
4. ASV time to spontaneous breathing
5. Episodes of nausea and vomiting, and use of rescue antiemetics
6. Length of ICU and hospital stay

**Figure 1.**
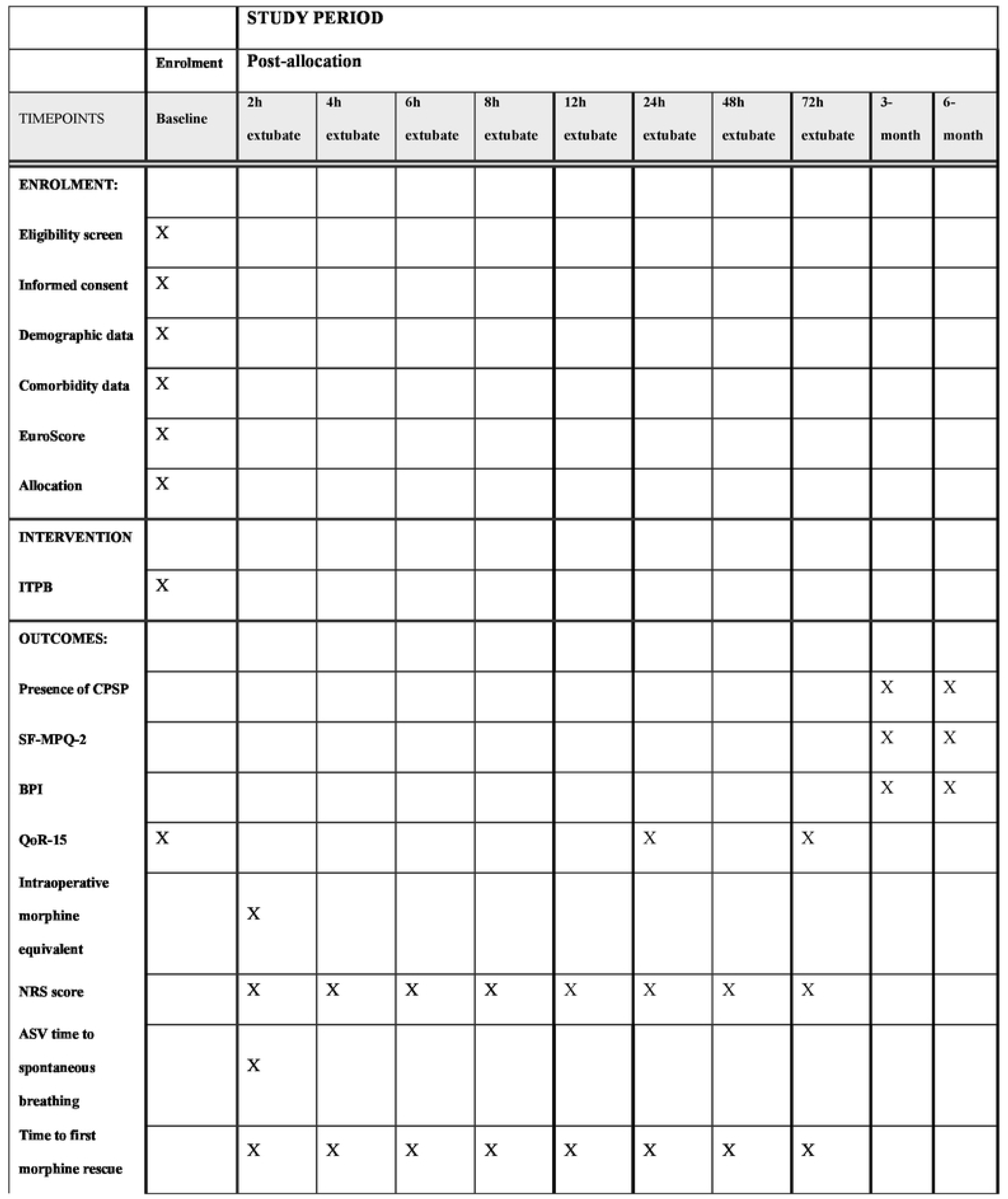
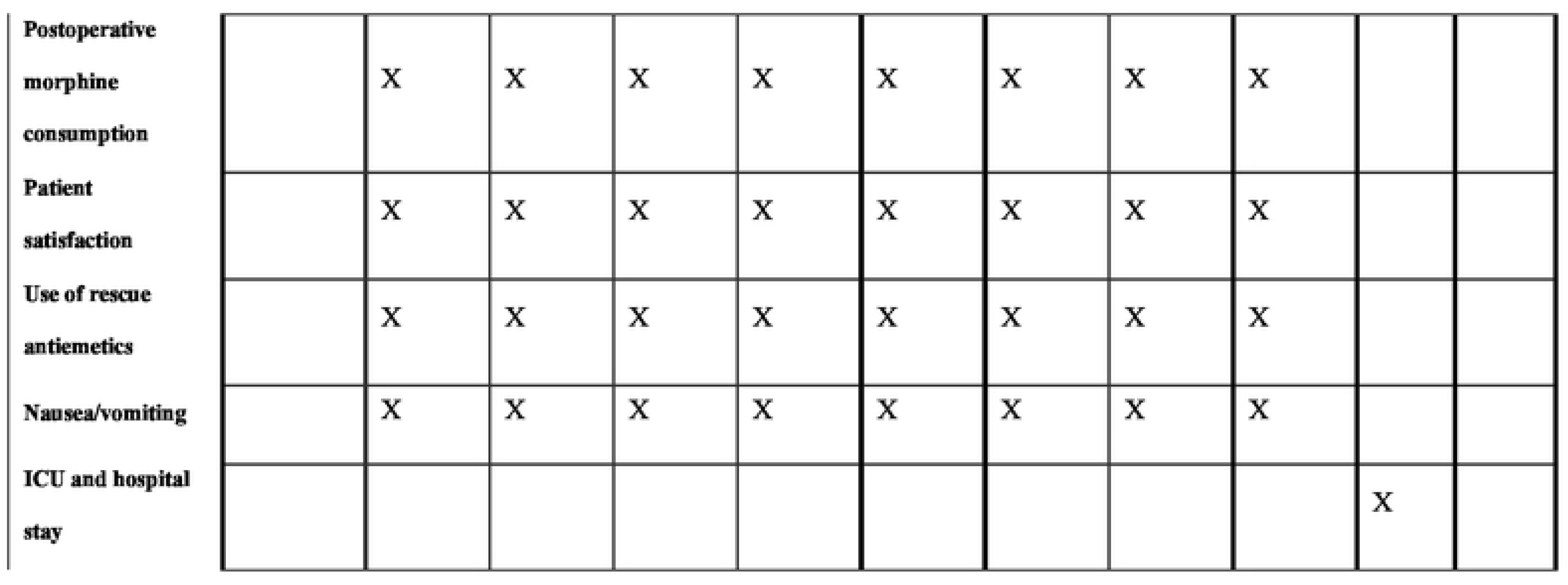
Assessments overview. NRS, numerical rating scale; ASV, adaptive support ventilation; QoR-15, Quality of Recovery questionnaire; BPI, Brief Pain Inventory; SF-MPQ-2, Short-Form McGill Pain Questionnaire

### Sample size calculation

Sample size was calculated using G*Power software version 3.1.9.3 (Kiel University, Kiel, Germany), based on the QoR-15 score at 24 hours postoperatively — the primary outcome. The minimum clinically important difference (MCID) for the QoR-15 score is eight points [38], and the typical standard deviation (SD) ranges from 10 to 16 [39–40]. Assuming a two-sided type I error of 0.05, type II error of 0.2, and a population variance of 144 (SD = 12), a sample size of 36 per group is required. Allowing for a 20% dropout rate as a result of loss to follow up after patients discharge, a total of 96 patients (48 patients per group) will provide 80% power to detect a mean difference of ≥8 points in the QoR-15 score at 24 hours between the two groups.

### Data analysis

All outcomes will be analyzed and reported on an intention-to-treat basis, with patients analyzed according to their randomized group regardless of protocol adherence. A secondary per protocol analysis will be conducted for patients who do not adhere fully to the study protocol. Given the repeated measures of pain scores over time, which are correlated, Generalised Estimating Equation (GEE) models will be used to assess the time effects of postoperative analgesia. Categorical data will be reported as counts and percentages. Continuous variables will be presented as mean (standard deviation) or median (interquartile range), depending on normality assessed using the Shapiro-Wilk’s test. Between-group comparison will be conducted using the independent sample *t*-test for parametric data and Mann-Whitney U test for nonparametric data. Categorical variables will be compared using the Chi-square test. Data analyses will be performed using SPSS 27.0 (IBM Corp, Armonk, NY), and GEE modelling will be conducted using Stata V.14 (Statam College Station, Texas, USA), with a Gaussian distribution, identify-link function, exchangeable correlation structure, and robust standard errors. A P-value of <0.05 will be considered statistically significant, without adjusting for multiple comparisons.

### Ethics, data management and dissemination

Patients will be screened for recruitment on the day prior to surgery, and the risks and benefits of the study will be explained. Written informed consent will be obtained from each patient. Participation is voluntary, and patients may withdraw from the study at any time without prejudice. Each participant will be assigned a unique identifier code for use throughout the study. All data will be entered into an electronic system by research team members trained in data entry. To ensure accurate, a second team member will verify the data. Data collection and study conduct will be monitored weekly by the research team to ensure consistent implementation of study protocols. All data will be kept confidential and stored on password-protected computers and in locked filing cabinets within the secure offices of the Department of Anesthesia and Intensive Care. Digital files will be securely deleted, and paper documents shredded, 5 years after the study. Only group-level data will be published, and access to individual data will be restricted to study investigators. Ethical approval for the project has been obtained from the Joint Chinese University of Hong Kong-New Territories East Cluster Clinical Research Ethics Committee. The study will adhere to local laws, the Declaration of Helsinki, the International Council for Harmonization of Technical Requirements for Pharmaceuticals for Human Use (ICH) Good Clinical Practice guidelines, and institutional policies. All adverse events associated with the study drug will be recorded by the research team and reported to the trial management committee. This committee, comprising external researchers and independent clinicians, will review all events within 48 hours and discuss them at regular trial committee meetings. The results of this study will be disseminated at international conferences and published in peer-reviewed journals.

## Discussion

Chronic postsurgical pain (CPSP) remains a significant and often debilitating complication following cardiac surgery, severely impacting patients’ daily functioning, health-related quality of life, and contributing to increased healthcare costs [1–4, 8–9]. Existing evidence highlights the crucial role of acute postoperative pain control in mitigating the development of CPSP. While various pain management strategies exist — including opioids, non-opioids, and multimodal analgesia — each has its limitations. Furthermore, current multimodal analgesia approaches, particularly for sternotomy pain, demonstrated limited efficacy [13]. Regional anesthesia techniques offer a promising avenue for enhanced pain control in cardiac surgery [14–17]. However, established neuraxial techniques such as epidural and intrathecal analgesia, despite their demonstrated efficacy, carry a potential risk of epidural haematoma in the context of systemic heparinization during cardiac surgery. Recent systematic reviews on paraspinal and chest wall blocks suggest potential benefits in reducing pain and opioid consumption, but are constrained by small sample sizes and methodological heterogeneity [16]. Techniques such as paravertebral blocks carry risks of pleural puncture and pneumothorax, while erector spinae plane block (ESPB) has shown inconsistent efficacy due to unpredictable local anesthetic spread [22–23]. Our own local experience with deep parasternal intercostal plane block also failed to demonstrate superior analgesia compared to conventional opioid-based approaches, despite a significant reduction in intraoperative opioid use [25]. This may be due to the limited duration of the block, which was insufficient to cover the prolonged surgical and postoperative recovery period. In addition, parasternal fascial plane blocks offer limited somatic (T2-T6) and visceral coverage.

This study addresses a critical research gap by investigating the efficacy of intertransverse process block, a novel regional technique, for pain management in cardiac surgical patients. To our knowledge, no existing data are available on the use of ITPB in this specific population. ITPB offers several potential advantages over existing techniques. Magnetic resonance imaging studies have demonstrated consistent spread of local anesthetic to the intercostal and paravertebral spaces, neural foramina, and epidural space, suggesting the potential for comprehensive somatic and visceral analgesia [30]. Its anatomical location, relatively distant from major vessels and the neuraxial space, may reduce the risks of complications such as epidural haematoma and pleural puncture associated with other paraspinal and neuraxial regional techniques. Furthermore, the closer proximity of ITPB to the neural foramina and epidural space may offer more consistent and reliable analgesia compared to techniques like ESPB. Our pilot in cardiac surgical patients has already demonstrated the safety and feasibility of ITPB, with improvements in Quality of Recovery scores at 24 hours post-extubation. Given patient-reported outcomes immediately after surgery is associated with the risk of developing CPSP, ITPB has a potential to reduce CPSP in this patient population. Ultimately, this trial will be crucial for optimizing pain management strategies and enhancing recovery after cardiac surgery, addressing a critical unmet need in patient care.

## Authors’ contribution

The protocol was designed and written by HMKW, and was critically reviewed by RKS, WTW, AKMC, ZHSY, PYC, STWT, MHMC, RHLW and KMH. All authors approved the final version of the manuscript.

## Data Availability

No datasets were generated or analysed during the current study. All relevant data from this study will be made available upon study completion.

## Acknowledgements

We would like to thank all the cardiac anesthesiologists, surgeons, nurses who contributed and participated in this study

## Supporting information

Supporting information checklist S1: SPIRIT checklist

Supporting information file S1: Project submitted to the ethics committee

Supporting information file S2: Opinions from the ethics committee

